# Application of Full Genome Analysis to Diagnose Rare Monogenic Disorders

**DOI:** 10.1101/2020.10.22.20216531

**Authors:** Joseph T. Shieh, Monica Penon-Portmann, Karen H.Y. Wong, Michal Levy-Sakin, Michelle Verghese, Anne Slavotinek, Renata C. Gallagher, Bryce A. Mendelsohn, Jessica Tenney, Daniah Beleford, Hazel Perry, Stephen K. Chow, Andrew G. Sharo, Steven E. Brenner, Zhongxia Qi, Jingwei Yu, Ophir D. Klein, David Martin, Pui-Yan Kwok, Dario Boffelli

## Abstract

Current genetic tests for rare diseases provide a diagnosis in only a modest proportion of cases. The Full Genome Analysis method, FGA, combines long-range assembly and whole-genome sequencing to detect small variants, structural variants with breakpoint resolution, and phasing. We built a variant prioritization pipeline and tested FGA’s utility for diagnosis of rare diseases in a clinical setting. FGA identified structural variants and small variants with an overall diagnostic yield of 40% (20 of 50 cases) and 35% in exome-negative cases (8 of 23 cases), 4 of these were structural variants. FGA detected and mapped structural variants that are missed by short reads, including non-coding duplication, and phased variants across long distances of more than 180kb. With the prioritization algorithm, longer DNA technologies could replace multiple tests for monogenic disorders and expand the range of variants detected. Our study suggests that genomes produced from technologies like FGA can improve variant detection and provide higher resolution genome maps for future application.

## Introduction

Current approaches to diagnosis of monogenic conditions include short-read sequencing of exomes or genomes ^1–6^ Although the diagnostic yield from these methods is promising, ranging from 26% to 40% ^1,5^, they leave many cases unresolved ^2,7^. The yield can be augmented by re-analysis against recently discovered disease-associated variants and genes, ^6,8^ or by using family-based analysis to identify *de novo* variants ^4^, but improvements are modest.

Two factors are principally responsible for diagnostic failures using short-read sequencing. First, short-read sequencing does not give a complete representation of the genome. For example, exome sequencing does not detect the majority of structural variants (SVs), cannot create chromosomal maps, misses variants in exons that are not captured efficiently, rarely detects repeats, and misses non-exonic variants ^9–13^. Even whole genome sequencing (WGS), which provides up to 9% additional diagnostic yield compared to exome sequencing ^5–7,14^ cannot detect all structural variants (especially duplications, inversions, and translocations), create chromosomal maps, or provide phasing information. In addition, detection of structural variants by short-read whole genome sequencing requires additional analytical processes that are often not fully implemented in clinical settings. ^15,16^ Second, genetic diagnosis of rare disorders often entails “experiments of one,” where many sequence variants found in the proband must be vetted against current knowledge (gene/variants and genome reference) to decide if variants meet the diagnostic criteria ^17^. Our incomplete biological knowledge limits our ability to identify the “causal variant” for any particular patient. Until we understand the functional consequences of more variants, or more patients with the same phenotypes are found, many candidate variants remain variants of uncertain significance.

Newly developed long DNA sequencing and mapping technologies can help solve the problem of incomplete genome analysis ^9,10,12^. Genome sequencing methods that produce long contigs (sets of adjacent DNA segments that together represent a consensus region of DNA) promise several advantages over short-read sequencing alone ^9–11,13,18^. First, long-read methods can resolve more easily large SVs (including large deletions, large insertions, translocations, and inversions), eliminating the need for additional genetic tests ^9,19^. Second, long-read sequencing detects insertions/deletions (indels) of intermediate size (500 bp to 50 kb) ^11^ more readily. These variants escape detection by clinical microarray because they are too small and by short-read sequencing because of clinical pipeline limitations or challenging filtering. Third, the methodology of genome reconstruction provides opportunities to detect rearrangement variants that have evaded detection ^11^. Fourth, single-basepair resolution of rearrangement breakpoints allows for determination of the precise location of each insertion or duplication, making it possible to see if structural variants disrupt genes or other sequences of functional significance. Finally, long contigs provide unique phasing information to determine the haplotype on which a variant occurs (e.g. *cis* or *trans*) and can help resolve recessive disease-associated alleles ^18^.

Here we test the diagnostic capabilities of an approach we call “Full Genome Analysis” (FGA), which combines linked-read sequencing technology and optical mapping to produce contigs with a median length of ∼100 Mb. To analyze the data in an unbiased and comprehensive fashion, an automated genetic variant interpretation pipeline was built to select and prioritize variants based on the individual patient phenotype. We show that FGA, when applied to patients with rare disorders in a clinical setting, leads to new diagnoses for patients with a variety of variant types. We further show that FGA is capable of detecting translocations, intermediate-sized copy number variants, phased biallelic variants – variations that are responsible for disease but usually missed by analysis of short-read sequencing data alone. Based on its efficiency in diagnosis, FGA opens the prospect of resolving more complex parts of the genome and identifying a more comprehensive set of genetic variants in rare disorder diagnosis ^9,10,12^.

## Results

### Automated Genetic Variant Interpretation Pipeline Performance

Using an automated genetic variant interpretation pipeline, we performed full genome analysis (FGA) on 50 undiagnosed cases to determine diagnostic yield and asked if it could help solve cases that had not been diagnosed with previous testing. The test was carried out in a clinical environment, allowing clinicians to propose unsolved cases for further genomic sequencing; cases were included only if prior testing was negative and characteristics of the case did not suggest any further specific test. Of the 50 cases, 23 had negative prior commercial trio whole exome sequencing and 42 prior negative microarray. FGA was performed and also compared to whole genome sequencing structural variant (SV) calling.

An automated pathogenicity assessment pipeline was built in-house to filter and prioritize variants based on the reported clinical features of recruited patients. In addition to analyzing single nucleotide variants (SNVs) and small indels, this pipeline was designed specifically to handle structural variations that often evade detection by short-read technologies. Our SV modules integrate the two complementary datasets (linked-read sequencing and optical mapping) and are capable of reporting clinically relevant translocations, inversions, deletions, duplications, insertions and other types of complex SVs of virtually all sizes. This pipeline ranks variants for every case and considers each potential inheritance pattern.

Overall, our automated pipeline identified 20 diagnostic cases (14 SNPs/indels and 6 SVs, **Table 1**). Most diagnostic cases (n=14) were ranked as the top variant in their respective inheritance/SV groups. By detecting additional variation beyond standard clinical testing modalities, FGA yielded novel genomic information for discovery in undiagnosed patients. The total diagnostic yield was 40% from the 50 cases tested (20 of 50 cases), with FGA detecting both new structural variants and single-nucleotide variants that were missed by previous sequencing and short read annotation. FGA diagnosed 35% of exome-negative cases (8 of 23 cases) (**Table 1**). Four of these were structural variants missed by exome sequencing, three were SNVs/Indels missed due to lack of annotation, and one was a suspected mosaic Indel, pending further validation. The diagnostic yield for cases that did not have prior exome was 44% (12 of 27 cases). We also identified candidate variants in another 60% (18 of 30 cases) for future follow-up.

### Full Genome Analysis

FGA solved three classes of cases where short-read sequencing or microarray analyses had previously failed to detect the causal variants: 1) cryptic heterozygous structural variants (e.g. *NHEJ1*-*IHH, WAC*), particularly variants of intermediate size; 2) translocations; and 3) missed phased heterozygous variants, for example in *trans* for recessive disorders (e.g. *TSPEAR*) (**Table 1**). Here we describe examples of diagnostic findings and the performance of the automated pipeline.

### Non-Coding Structural Variation

In a <18-year-old female with craniosynostosis and syndactyly, we found a rare 32 kb heterozygous *de novo* intronic duplication within the *NHEJ1* gene (case 1703). FGA identified the breakpoints of a duplication at chr2: 219,102,933 - 219,134,970 (genome version GRCh38) (**Figure 1**). Only by familial mapping studies have similar duplications been described in cases of craniosynostosis and syndactyly ^20,21^ (named Chromosome 2q35 Duplication Syndrome, OMIM #185900). The breakpoints identified here narrow the critical region of the *NHEJ1* intron that is important for the condition ^20,21^. This heterozygous *de novo* duplication was detected by both optical mapping (Bionano) and linked-read sequencing (10x Genomics) technologies. The duplication occurred adjacent to the original segment in tandem, information readily identified using optical mapping (**Figure 1, Panel A**). This mid-sized structural variant (∼32 kb) was not detected by standard microarray analysis because it is small and intronic. It also escaped detection by our short-read whole genome sequencing (WGS) copy number variant calling–but was easily identified by FGA (**Table 2**).

**Figure 1.**
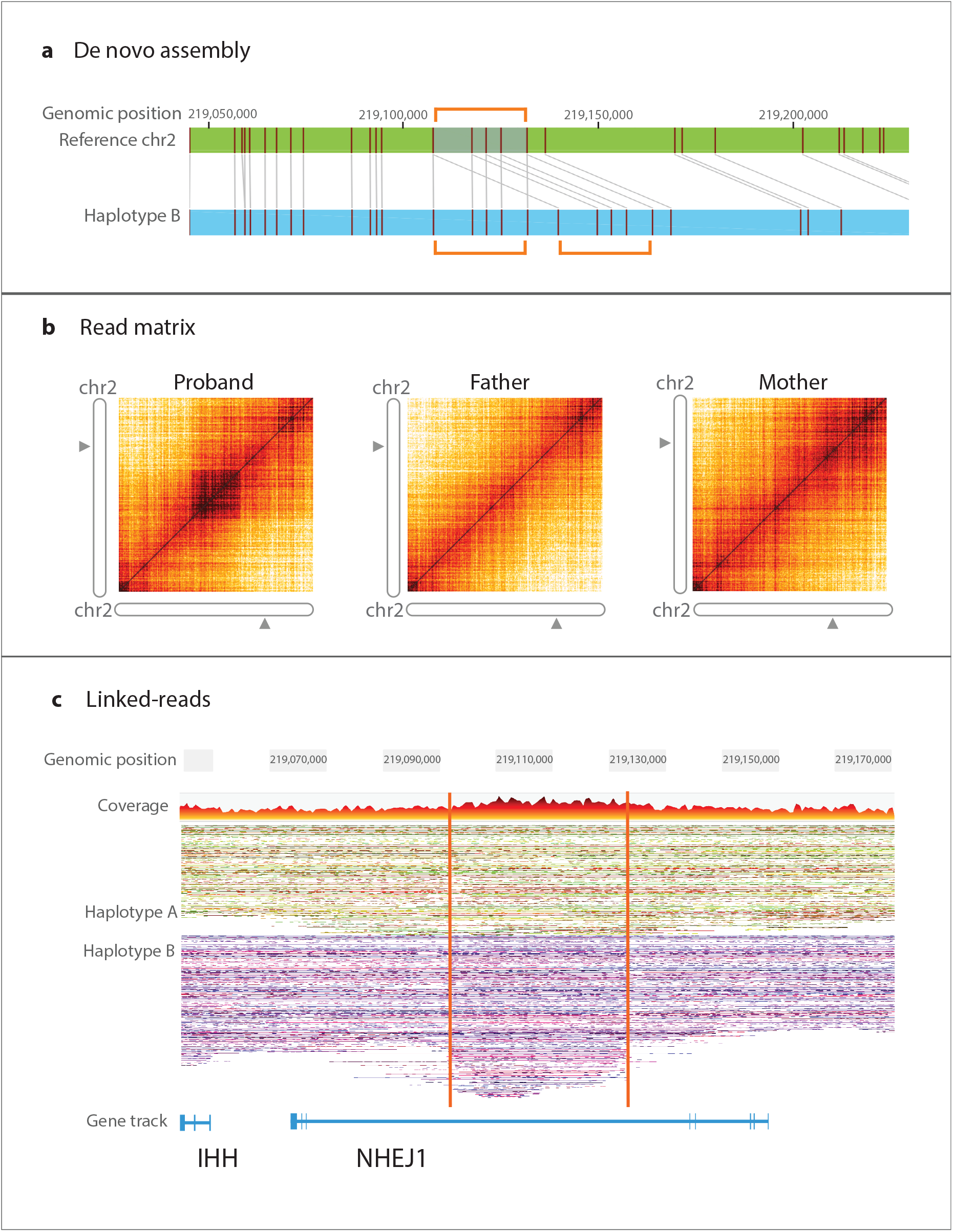
Heterozygous, intronic tandem duplication (32 kb) in *NHEJ1*. The region affected (chr2: 219,102,933 - 219,134,970, 2q35, genome version GRCh38) corresponds to an *IHH* upstream enhancer and narrows the diagnostic interval for this condition. **Panel A:** depicts a *de novo* assembly (light blue) and its alignment to reference (green). The labeled motifs in the reference genome (vertical maroon lines) are duplicated in Haplotype B and their orientation demonstrates the duplication occurred adjacent to the original sequence, in tandem. **Panel B:** shows a matrix view of linked reads. The dark orange square in the left panel (proband), illustrates a higher density of barcode overlap in the read matrix compared to parents, indicating the variant likely occurred *de novo*. **Panel C:** contains phased haplotypes generated using linked-read data. Haplotype B, in purple, contains the intronic region with higher number of linked-reads due to sequence duplication.

The duplication affects an enhancer for the Indian Hedgehog (*IHH*) gene, located upstream in the third intron of the neighboring *NHEJ1* ^20^. ENCODE data support the enhancer function of this intronic region. The structural variant breakpoints defined by FGA narrow the intronic region responsible for this condition.

### Genomic Rearrangements

It can be challenging to readily detect translocations with current clinical sequencing pipelines without specific additional informatic analyses. In contrast, we were able to detect translocations readily using FGA with our automated pipeline. For example, we found a germline translocation between chromosomes 1 and 9 in a <18-year-old male with a history of neuroblastoma and developmental delay who had negative microarray and exome sequencing (**Figure 2** – both linked-read genome sequencing and optical mapping support the translocations, case 0703). Trio analysis indicated that the translocation occurred *de novo*; it was subsequently verified by cytogenetic chromosome analysis (Karyotype: 46,XY,t(1;9)(p32.3;p21). FGA identified the precise breakpoint locations on chromosome 1 and chromosome 9 (chr1: 49,553,194 and chr9: 29,096,674, respectively, genome version GRCh38). The breakpoints were non-exonic, occurring in an intronic region of *AGBL4* and an upstream/untranslated region near *LINGO2*. FGA also revealed that the translocations occurred on the paternal allele, with a small breakpoint deletion suggesting non-homologous end joining, with an additional maternally inherited intronic deletion present on the other allele. *AGBL4*, encoding a cytosolic carboxypeptidase, has a potential role in neuroblastoma, autism and developmental delay. Copy number alteration has also been reported in *LINGO2* in neuroblastoma cell lines ^22,23^. Although several other genes in this case had *de novo* variants (*MYH11, GABRA2, TFE3*), none of these were clearly etiologic.

**Figure 2.**
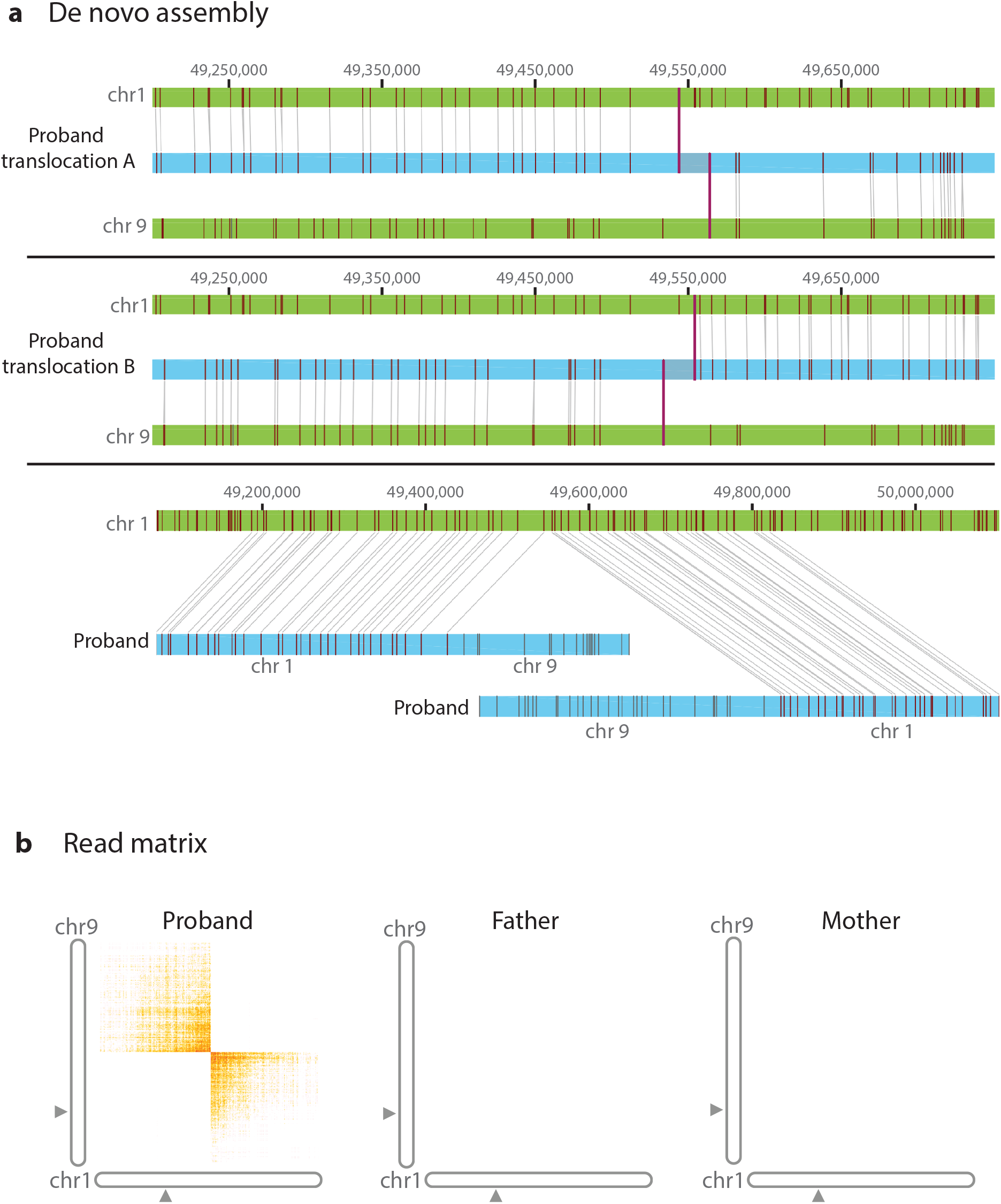
Structural rearrangement detection with *de novo* assembly and linked reads; t(1:9)(p33,p21). **Panel A** contains *de novo* assemblies of chromosome 9 and 1. Genomic coordinates in grey at the top and the reference assembly in green (reference GRCh38). The proband assembly map is shown in blue with vertical maroon lines that show matching label patterns. The first and second panels show two *de novo* assembly maps that align to reference chromosomes 1 and 9 and the translocation breakpoint where the alignment switches. The third panel depicts two assembly maps in chromosome 1 with segments that align and segments that do not align to the reference due to the translocation. **Panel B** shows the matrix view of linked reads that contains unexpected barcode overlap (in orange) between chromosome 1 and 9, corresponding to the intronic point of fusion between the two. This overlap is absent in the parents.

Neuroblastoma predisposition genes *ALK* and *PHOX2B* were also negative ^24^. The *de novo* translocation suggested a new etiology for this condition, which could be explored in future studies ^22,25^. Interestingly, the short read WGS copy number calling shows hundreds of potential breakpoint junctions that needed further analysis for diagnostic use. In contrast, FGA had at least 8-fold fewer candidates. Furthermore, *de novo* assembly from FGA promptly identified the event as a translocation.

Translocations have implications for future reproductive risks. A diagnostic strategy that encompasses what chromosome analysis and microarray do in a single diagnostic test could also serve to detect balanced events which are important for family planning in carriers. More complex rearrangements were also promptly detected among significantly fewer candidates and localized with FGA (e.g. unbalanced insertional translocation) providing novel variants for future characterization.

### Deletions

FGA was also capable of pinpointing genomic breakpoints of clinically significant deletion copy number variants. FGA identified 36kb deletions disrupting *TANGO2* (OMIM #616878, case 5103) in siblings with a history of episodic rhabdomyolysis, metabolic acidosis, and ketosis (**Figure 3**). A 1480 bp *de novo* deletion in *WAC* (Desanto-Shinawi syndrome, OMIM #616708 case 4203) was found in a male patient with seizures, hypotonia, developmental delay and nonfamilial features like low-set ears and brachycephaly. We also identified a 5000 bp *de novo* deletion in 2p15 in a female with seizures, developmental delay (2p16.1-p15 deletion syndrome, OMIM #612513, case 4803) which implicates *USP34*. FGA simultaneously identified and verified such variants and breakpoints without additional copy number prediction tools or external validation required by current short-read sequencing pipelines. Indeed, short-read sequencing copy number calling was able to detect these deletions albeit with sometimes incorrect zygosity. FGA had an advantage compared to short-read sequencing in identifying deletions in challenging regions of the genome near segmental duplications, in agreement with our previous studies ^9^.

**Figure 3.**
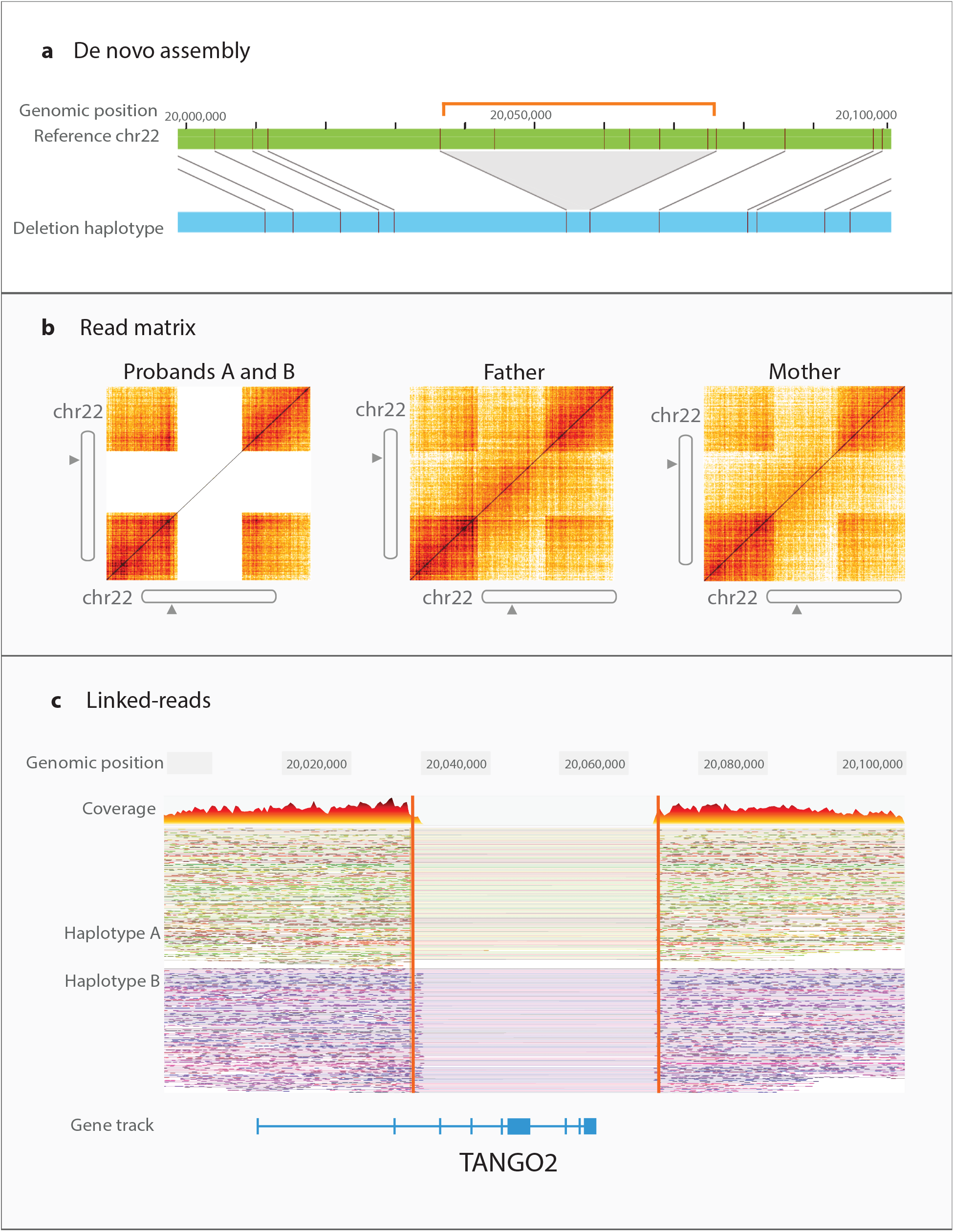
Deletion disrupting *TANGO2*. (chr22: 20,039,637 – 20,075,714 and chr22: 20,041,469 – 20,075,432, genome version GRCh38). **Panel A:** *De novo* assembly (light blue) demonstrates missing sequence labels with respect to reference (green). The orange bracket and gray triangle shows the deleted region. **Panel B:** Matrix view with absent signal from intervening region demonstrates proband with biallelic deletion. **Panel C:** Deletion is also seen by drop in coverage in both haplotypes in linked-read data.

### Small Variant Detection and Biallelic Phased Variants

FGA also yielded coding variants, similar to short-read WGS, however phasing was now uniquely possible given the longer DNA segments. Discerning that variants reside on separate chromosomes is important for diagnoses involving compound heterozygous recessive variants; FGA is capable of making this determination in a single proband test. We identified two *TSPEAR* variants in a female with oligo/hypodontia, missing 15 adult teeth, but no previous family history. The two *TSPEAR* NM_144991 variants were found 180 kb away from each other. FGA phasing clearly showed that the variants occurred in *trans*, suggesting that both parents are heterozygous carriers, which was confirmed. The first variant is a 10 base insertion which leads to frameshift, c.51_52insGGCCCCCGGC, p.His18fs, while the second variant is nonsense, c.1281G>A, p.Trp427Ter; together the variants confirm the biallelic etiology (**Figure 4, Panel A**). The large phasing block (chr21: 29,801,272 - 44,927,448, genome version GRCh38) created by FGA was able to discern the two haplotypes and determine that the variants are in *trans*, even with the affected individual’s sequence only. *TSPEAR* has recently been associated with tooth agenesis, thus missed by prior sequencing, and loss-of-function variants in *TSPEAR* are associated with ectodermal dysplasia 14, hair/tooth type, with or without hypohidrosis (OMIM #618180) ^26,27^. Indeed, WGS may have identified these variants, but only phasing using FGA is sufficient to make a diagnosis on proband alone. In detecting compound heterozygous variants, phasing information is valuable since one of the variants might be *de novo* and data from parents are not always available to exclude the possibility that variants are *in cis*. FGA also localized additional diagnostic *de novo* single nucleotide variants, similar to whole genome or exome sequencing, but also determined which parental allele was affected by the mutation. For example, a <18-year-old girl with short stature, neurodevelopmental disability, and cardiac valvular disease who had not had prior exome testing had a *de novo* missense variant by FGA, occurring on the paternal allele in *SMAD4*, NM_005359:c.1498A>G, p.Ile500Val (**Figure 4, Panel B**), diagnostic of Myhre syndrome (OMIM #139210), which is associated with increased risk of pericardial, pulmonary, and tracheal fibrosis, as well as skeletal and vascular complications. The diagnosis was useful for patient management. *De novo* mutations become more common with advancing paternal age ^28^. For some medical conditions, the allele affected (maternal or paternal) could also determine whether mutations are clinically significant or not (e.g. imprinted regions).

**Figure 4.**
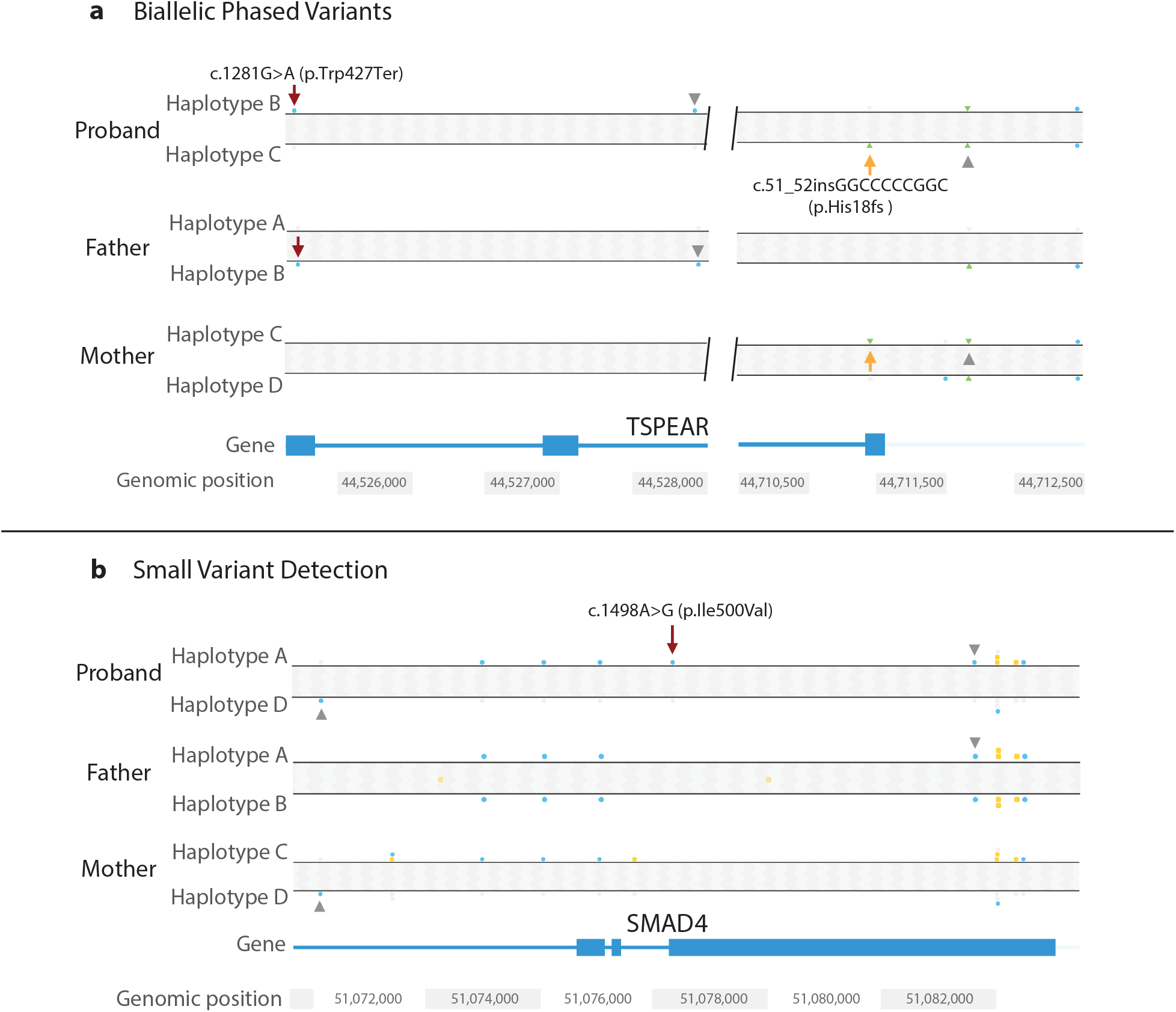
Variant haplotype distinction. **Panel A** shows compound heterozygous *TSPEAR* variants (NM_144991). Phasing was successful for etiologic variants 184,756 bp apart given a phasing block of 15.1 Mb, which is not possible with short read sequencing. Maroon and yellow arrows point to each variant. Grey arrowheads point to single nucleotide polymorphisms that confirm *trans* orientation in relation to parental haplotypes. **Panel B** shows a *de novo SMAD4* pathogenic variant (NM_005359) identified by linked-read sequencing, also detectable by short read sequencing. Haplotype analysis showed the variant occurred on the paternally-inherited allele, Haplotype A. Variant position is indicated with a maroon arrow. Grey arrowheads point to

## Discussion

In genomic medicine, rare disease diagnostics has traditionally been challenged by the rarity of the disorders and testing limitations. Here, we described the FGA approach with automated analysis using linked-read sequencing and optical mapping to evaluate a full spectrum of genetic variants implicated in rare genetic diseases. The automated pipeline integrates the longer DNA technologies into the diagnostic realm by enabling a streamlined variant detection protocol and minimizing biases introduced during the analysis process. This data-driven approach results in a drastic decrease in human intervention and ensures that every case is evaluated thoroughly. We find that genome assemblies can be used in clinical testing strategies detecting all types of genetic variants concurrently. FGA detects and localizes SV such as duplications that are missed by WGS and can easily identify translocations and phase variants across long distances. With variant detection from longer DNA technologies, we can improve detection of diagnostic variants and provide higher resolution genome maps for future studies.

For individuals with undiagnosed conditions, these technologies encompass what is currently provided by the combination of chromosome analysis–karyotyping, microarray testing, and short-read WGS ^5^. By identifying novel SVs and phasing, it provides diagnostic information beyond current clinical tests. The automated pipeline also provides internal validation of SVs, bypassing the need for additional time and blood for testing ^14^. By constructing *de novo* genome assemblies and identifying variants that do not map to the genome reference, these technologies can also provide additional information for future analysis. These strengths make the technologies highly suitable for early implementation in diagnostic evaluations, particularly if a specific genetic condition or type of variant is not immediately suspected ^5^.

As expected, genome assemblies are able to detect duplications and translocations more efficiently than the short-read sequencing. The longer DNA techniques also have practical advantages over traditional genetic testing strategies because they can detect phased variants for recessive conditions, as well as the full spectrum of structural variants. Therefore, FGA makes it possible to effectively test probands even when parents/family members are not available for testing. This is useful in intensive care units or in other settings where rapid diagnosis is vital to clinical care ^29,30^. Variant phasing or the *cis* or *trans* configuration can be critical in the rapid evaluation for clinical significance. FGA also returns a high-quality genome reconstruction, which is useful for resolving complex or novel regions of the genome. Such *de novo* assemblies are not reference-dependent and SV calling can be achieved without making inferences that are necessary in short-read sequencing ^9–11,19^.

The number of diagnostic cases attributable to SVs was striking in our study, as 50% of exome negative cases (4 out of 8 cases) were solved by identifying an SV or rearrangement. We also identified at least one highly probable SV or SNV candidate in more than half of the remaining undiagnosed patients. These cases do not meet diagnostic criteria due to several reasons. SVs overlapping similar regions do not always produce the same phenotype. This is particularly limiting since most SVs are not recurrent and thus do not share identical breakpoints. Furthermore, unless a critical region can be established or a syndrome is associated with very distinctive phenotype, it is unclear whether an SV or SNV can be diagnostic even if it is *de novo*. Most importantly, SV/CNV databases are strikingly sparse and inconsistent, in contrast to SNV databases. Further genotype correlation and functional testing are needed in the future.

The application of hybrid technologies with long-range sequencing, like FGA, in genomic medicine is not without limitations. First, our automated variant interpretation pipeline is based on existing annotation databases. Genetic variations cannot be ranked or annotated well if they are not found in these resources ^14^. Second, even with the use of long molecules averaging 200-300kb in our optical mapping experiments, they are not long enough to resolve the large, near-identical segmental duplications in some of the most complex regions of the human genome. A small number of these complex regions remain inaccessible despite using long-range sequencing and mapping technologies ^9^. Truly whole or complete sequencing of genomes depends on the technical platform, analytical pipelines and thorough annotation ^5^. Third, the current human reference genome is a set of composite haplotypes generated from 8 anonymous DNA donors ^31^. As such, there are functionally important sequences found in many people around the world but that are missing from the reference genome ^10,32^. Since the reference genome serves as the benchmark for all analyses, missing sequences are never assessed, thus making variants in these regions undiagnosable.

FGA can be implemented for diagnostic purposes with minor modification of workflow in the clinical laboratory. Sample handling must be adapted to protect DNA integrity, which is required to obtain longer DNA fragments. The bioinformatic workflow is easily implemented in a clinical setting with phased haplotypes and structural variants as direct output. This is in sharp contrast to the workflow required for the detection of structural variants from short-read data.

We can expect that whole-genome sequencing is becoming the method of choice for genetic diagnosis, given the greater number of variants relative to exome sequencing or microarray analysis ^5,6,14^ In choosing technology for the acquisition of whole-genome data, one should consider costs and the complexity of analysis, as well as the completeness of the data and the continuing value of the data for future reanalysis. The inherent amount of missing data in genomes generated by short-read sequencing reduces their ability to complete clinical diagnoses in challenging cases. Data reanalysis is becoming a successful strategy to identify variants that underlie disease in a patient’s genome ^14^; as our understanding of deleterious variants grows, it is possible to revisit previously acquired data and assign significance to previously detected variants. FGA’s ability to acquire a more extensive set of variants increases the likelihood that future reanalysis will be productive. More importantly, by identifying previously unknown variants, FGA makes it possible to explore their functional significance.

The increase in diagnostic yield produced by FGA in this study, attributable to advances such as structural variant detection, has made it possible to solve cases that were negative by short-read sequencing. Full realization of FGA’s potential to provide comprehensive detection of clinical variants will require a combination of automated capture of phenotypic terms with expanded expertise in variant interpretation ^14^. Comprehensive assessment of the genome in every undiagnosed patient would rapidly produce both genome maps of annotated functional variants and new diagnostic possibilities. The result would be a better understanding of population variation, and improved diagnostics for direct clinical care.

## Methods

### DNA extraction and preparation

High molecular-weight DNA was extracted and isolated using the Bionano Prep Blood Isolation Kit following the manufacturer protocol (Bionano Genomics). Bionano optical mapping libraries were prepared following the manufacturer protocol (Bionano Genomics). 10x Genomics linked-read sequencing libraries were built as published^9^ using the GemCode platform (10x Genomics).

### Optical mapping and linked-read data generation and processing

Optical mapping on the Bionano Irys and Saphyr platforms was used to produce *de novo* assemblies and identify structural variants and rearrangements. DNA was labeled using Nick, Label, Repair and Stain (NLRS) and/or Direct Label and Staining Technologies (DLS). The first uses a nicking endonuclease that recognizes a specific 6-7 basepair sequence and creates a single-strand nick, filled with fluorescent nucleotides. The second uses a single direct-labeling enzymatic reaction to attach a fluorophore to a specific 6-basepair DNA sequence motif. Labeled DNA libraries were loaded onto the Bionano Genomics Irys™ Chip or Saphyr™ Chip, linearized and visualized using the Irys™ or Saphyr™ system, which detects the fluorescent labels along each molecule. Single molecule maps were assembled *de novo* into genome maps using Bionano Solve with the default settings ^12^. Genome assembly and alignment was performed using IrysView/IrysSolve software. For optical mapping, we performed embedding of cells, long DNA extraction and Chip run over a total 3.25 days.

Sequencing data was obtained from 10x Genomics linked-read libraries sequenced to ∼60X coverage using an Illumina sequencer. Reads were aligned to GRCh38 using LongRanger and variants were identified using the callers integrated in the 10x pipeline including GATK Haplotype caller for SNPs and indels. SNPs and indels were kept for analysis if the minor allele frequency is ≤ 5% as reported in the gnomAD database.

### Automated variant interpretation pipeline

An automated clinical interpretation and prioritization pipeline was built in-house using custom and publicly available software. Electronic health records were exported into JSON format for parsing with clinical natural language processing (NLP) algorithm ClinPhen. Since some HPO terms were already manually curated from previous sequencing studies, these terms were combined with the non-redundant ones generated from NLP. HPO hierarchical terms separated by 1 degree were also included as part of the clinical phenome. Every HPO term (*h)* is assigned a weight, which is defined as the inverse of the total number of disease genes associated with it.

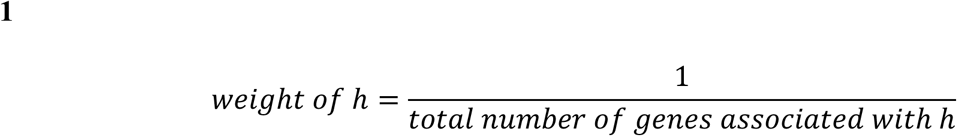

Next, we overlapped the clinical phenome of the proband with a list of known phenotypic features associated with mutations in a given gene (*G*). The overlapping terms were used to calculate a gene sum score to identify and rank clinically relevant genes.

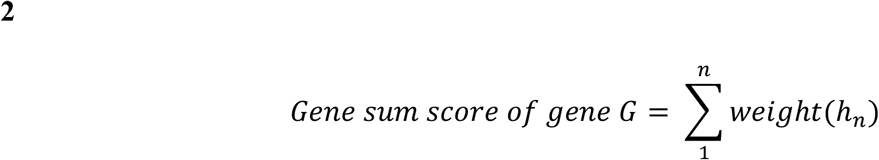

SNP and indel exonic variants identified from the proband were overlapped with the ranked gene list. The same strategy was applied to all SVs overlapping genic exons. Scores were normalized for comparing and calculating confidence scores. All SVs were vetted against a set of regions known to be associated with deletion and duplication syndromes. Structural variants and copy number variants were screened first using the entire cohort, for single or double occurrences. These were then compared to the gnomAD SV database. Of note, three out of the four diagnostic SVs are absent from gnomAD and were also rare in our platform. Additionally, all translocations and inversions were included by default. Prioritized variants were reviewed manually to determine which one was diagnostic. We initially focus on structural variants and did not systematically annotate deep intronic variation or short tandem repeats. Mitochondrial DNA candidates were annotated and manually verified.

This tool parses SNPs, indels, and structural variations (SVs) from 10x Genomics linked-read and Bionano optical mapping data based on trio sequencing (singleton is allowed). SNPs/indels analysis can be done alone or in combination with SV analysis. In general, this tool parses a patient’s electronic health record in JSON format and outputs a clinically relevant gene list. This gene list is then used to inform how genetic variants are prioritized. Genetic variants (SNPs, indels, and SVs) are vetted against a set of controls and parents. For SNPs and indels, variants are filtered based on allele frequencies reported by gnomAD. Small variants reported as likely benign or benign by either Clinvar or Intervar are discarded from the pipeline. For SVs, the prevalent of these variants are compared against a set of 1KGP + CIAPM control sequenced previously by the Kwok lab. See below for more details.

A pre-processing step (for SNPs and indels) is required to run this software. This pre-processing step takes the 10xG GATK output and applies filters based on GQ, DP, and PASS. This step removes the bulk of the variants that are likely to be artifacts. Remaining variants are annotated using Intervar, which is a wrapper for Annovar and it assigns ACMG pathogenicity to all variants. Variants are additionally filtered for frameshift, nonframeshift, nonsynonymous, stopgain, stoploss, and splicing. They are overlapped with the ranked gene list generated previously. All remaining variants are ranked by the reported pathogenicity based on ClinVar/Intervar and then by the gene sum score (see manuscript for details).

For SVs, insertions, deletions, and duplications are annotated with known exons (exon-level not gene-level). Duplications and deletions are additionally used to search for known microdeletion and microduplication syndromes. Inversions and translocations are annotated with known genes (gene-level) and every call in these two categories are always reported.

SV scripts have been designed to analyze BioNano Optical Mapping Data (.smap file format) and 10x Linked Reads Data (.vcf file format). All SV scripts read a proband file, mother file, father file, and a reference file which consists of SV calls from the 1000 Genome Project cohort as well as SV calls from all other parents in the study other than the parents of the proband being analyzed. The scripts output filtered proband calls with additional descriptor columns as a tab-delimited txt file.

BioNanoDeletions, BioNanoInsertions, and BioNanoDuplications select calls of the SV type and eliminate calls below the inputted confidence threshold (default: 0.5). They perform a 50% reciprocal overlap with the reference file and remove calls that overlap. They perform a 50% reciprocal overlap with the inputted mother and father file separately and append columns (Found_in_Mother, Found_in_Father) to describe the overlap (True/False). They overlap with exons and phenotypes and append columns (Gene, Phenotype) with the gene name and phenotype if found.

BioNanoInversions and BioNanoTranslocations select calls of the SV type and do not filter for confidence. They create 20kb intervals around the start point and end point of the call. They overlap the start and end intervals with the reference file and remove calls that overlap.

They overlap the start and end intervals with the mother and father file separately and append columns (Found_in_Mother, Found_in_Father) to describe the overlap (True/False). They overlap start and end intervals with genes and phenotypes and append columns (Gene, Phenotype for start point; Gene2, Phenotype2 for end point) with the gene name and phenotype if found.

tenxDeletions reads the 10x Deletion calls (“dels.vcf”) and performs a 50% reciprocal overlap with reference file and removes calls that overlap. It performs a 50% reciprocal overlap with the inputted mother and father file separately and appends columns (Found_in_Mother, Found_in_Father) to describe the overlap (True/False). It overlaps with exons and phenotypes and appends columns (Gene, Phenotype) with the gene name and phenotype if found.

tenxLargeSvDeletions and tenxLargeSvDuplications read the 10x Large SV calls (“large_svs.vcf”) and select calls of the SV type. They perform a 50% reciprocal overlap with reference file and remove calls that overlap. They perform a 50% reciprocal overlap with the inputted mother and father file separately and append columns (Found_in_Mother, Found_in_Father) to describe the overlap (True/False). They overlap with exons and phenotypes and append columns (Gene, Phenotype) with the gene name and phenotype if found.

tenxLargeSvInversions, tenxLargeSvUnknown, and tenxLargeSvBreakends read the 10x Large SV calls (“large_svs.vcf”) and select calls of the SV type. They create 10kb intervals around the start point and end point of the call. They overlap the start and end intervals with reference file and remove calls that overlap. They overlap the start and end intervals with the mother and father file separately and append columns (Found_in_Mother, Found_in_Father) to describe overlap (True/False). They overlap start and end intervals with genes and phenotypes and append columns (Gene, Phenotype for start point; Gene2, Phenotype2 for end point) with the gene name and phenotype if one is found. For unknown and breakend types, only variants with quality score > 1 standard deviation above the mean are reported. All reported coordinates are based on hg38.

### Comparison of short-read whole genome sequencing to the genome assembly technologies

To assess structural variant calls, we removed linked-read barcodes from sequencing reads to generate short-reads and performed short-read whole genome calling copy number using Manta with default settings ^33,34^. From Manta output we assessed deletions, duplications, and breakends called using the short-read data and compared these to output from 10x linked reads and Bionano optical mapping, with attention to variant size, zygosity and type of variant called. We also identified if calls passed more stringent high-quality filters in each of the three platforms.

### Approvals and Phenotypic Assessment

The study was approved by the Institutional review board of Children’s Hospital Oakland and University of California, San Francisco (UCSF), Committee for Human Subjects Research. Recrui™ent was from UCSF Benioff Children’s Hospital Medical Genetics and Genomics clinics. We focused on cases of two types, chosen to demonstrate the capability of FGA: cases in which whole-exome sequencing had not returned a causal variant; sporadic cases from the pediatric population that are suspected to have a genetic basis, but fall into no clear syndrome and have no clear candidate target for conventional genetic diagnosis. Individuals with undiagnosed conditions and unaffected parents were offered testing and underwent an informed consent process prior to blood draw. The nature and possible risks of the study were explained in the consent process. Phenotypic evaluation was performed by clinical review by at least two genetics professionals, and human phenotype ontology terms were curated for each case.

## Data Availability

Sequence libraries available upon request.

## Acknowledgements

Funded by the California Initiative to Advance Precision Medicine to D.M. the Marcus Program in Precision Medicine and Established Investigator Grants to J.T.S. and P-Y.K., and the National Human Genome Research Institute of the National Institutes of Health under award R01 HG005946 to P-Y.K. The National Science Foundation Graduate Research Fellowship under Grant No. DGE 1752814 support for A.G.S. We thank the families for their participation. We thank the California Initiative to Advance Precision Medicine and the California Governor’s Precision Medicine Advisory Committee.

## Author Contributions

J.T.S., D.M., P-Y.K., D.B. conceived and designed the study. J.T.S., A.S., R.C.G., B.A.M., J.T., D.B., H.P., Z.Q., J.Y., O.K., D.M., P-Y.K., D.B. acquired data. J.T.S., M.P-P., K.H.Y.W., M.L-S., M.V., H.P., S.K.C. analyzed data. J.T.S., M.P-P., K.H.Y.W., M.L-S., A.S., R.C.G., B.A.M., J.T., D.B., H.P., A.G.S., S.E.B, Z.Q., J.Y., O.K., D.M., P-Y.K., D.B. interpreted data. M.P-P., K.H.Y.W., M.V., S.K.C. created software used in the work. J.T.S., M.P-P, K.H.Y.W., D.M., P-Y.K., D.B. drafted the manuscript. J.T.S., M.P-P., K.H.Y.W., M.L-S., A.S., R.C.G., B.A.M., J.T., D.B., H.P., A.G.S., Z.Q., J.Y., O.K., D.M., P-Y.K., D.B. provided critical revisions. S.E.B. was unable to review the final manuscript due to injury. All authors contributed to the manuscript. J.T.S., M.P-P., K.H.Y.W. contributed equally to this work.

## Other Author Information

M.L-S. present affiliation: Dovetail Genomics. K.H.Y.W. present affiliation: Color. B.A.M. current affiliation: Kaiser Permanante.

## Data Availability

Sequence libraries available upon request.

## Code availability

The automated variant interpretation pipeline is hosted on GitHub (https://github.com/wongkarenhy/Full-Genome-Analysis-Pipeline).

